# Factors associated with Alzheimer’s Disease Dementia prevalence in the United States: A county-level spatial machine learning analysis

**DOI:** 10.1101/2024.07.16.24310529

**Authors:** Abolfazl Mollalo, George Grekousis, Andreana Benitez, Hermes Florez, Brian Neelon, Leslie A. Lenert, Alexander V. Alekseyenko

## Abstract

**Background:** In recent years, a growing body of literature has examined the impact of neighborhood characteristics on Alzheimer’s disease (AD) dementia. However, spatial variability of the most influential variables and their relative importance to AD dementia prevalence remain underexplored.

**Methods:** We compiled various widely recognized factors to examine spatial heterogeneity and associations with AD dementia prevalence utilizing non-linear geographically weighted random forest approach.

**Results:** The model outperformed conventional ones, with an out-of-bag R² of 74.8%. Key findings showed the normalized difference vegetation index as the most influential environmental factor in 15.1% of US counties, lack of leisure time physical activity in 12.7%, binge drinking in 9.1%, and mobile homes as the main socioeconomic factor in 13.3% of US counties.

**Discussion:** Spatial machine learning analyses suggest that AD dementia prevalence could be impacted by county-specific targeted interventions that improve green space, reduce air pollution and support greater access to physical activity.

## BACKGROUND

While there is currently no approved treatment for AD dementia [1], neighborhood environments where persons living with AD dementia reside can significantly impact the progression of the disease [2-5]. For example, an age-friendly environment has been shown to be crucial for older adults with limited mobility who often spend more time in their immediate geographic surroundings [6]. Further, evidence suggests that higher neighborhood deprivation is linked to lower cognitive function and increased depressive symptoms [7, 8]. Mechanistically, long-term exposure to air pollution and, specifically, fine particulate matter (PM2.5) can trigger brain inflammation and increase the risk of cognitive decline and neuroinflammation [9-11]. Conversely, greater exposure to green spaces can reduce stress and mental fatigue, promote physical activity, and lower the risk of cognitive impairment [12-16]. Access to healthcare and community resources is crucial for managing the disease and alleviating caregiver burden [17].

Geospatial analysis and modeling can provide a robust framework for examining the impact of neighboring environments on the geospatial distribution of AD dementia [18]. In the US, it has been used for the identification of spatial clusters of AD mortality rates [19], spatiotemporal clusters of dementia mortality [20], and spatial and spatio-temporal hotspots of dementia hospitalization rates [21]. However, only a limited number of geospatial modeling studies have been conducted to examine the local impact of neighborhood characteristics on AD dementia distribution. Karway et al. (2024) analyzed Medicare beneficiaries’ data using geospatial methods to assess the impact of cardiometabolic diseases (CMD), smoking, and educational attainment on the incidence of AD. They computed county-level AD incidence, adjusted for age, sex, and race using the spatial Bayesian latent Gaussian approach. They found strong agreements between the spatial patterns of AD and CMD, moderate association with smoking, and a weak (inverse) relationship with educational attainment [22]. Using the same approach, Alhasan et al. (2024) investigated the link between neighborhood attributes and AD dementia incidence in South Carolina. They found higher AD dementia incidence associated with poverty, air pollution, rural settings, and limited access to nutritious food [23].

Additionally, a limited number of geospatial models have examined the impact of socioeconomic, environmental, and lifestyle characteristics on AD dementia distribution. One widely used model to assess the regional impact of these variables on various health outcomes is geographically weighted regression (GWR) [24, 25], which relies on several challenging assumptions, such as local linearity between outcome and predictors [26], and is sensitive to outliers and multicollinearities [27]. In contrast, random forest (RF), a nonlinear and non-parametric decision tree-based model, is not based on statistical assumptions and is less prone to overfitting and outliers [28, 29]. Spatial variants of RF have been previously applied to model COVID-19 [27], hypertension [30], malaria [31], asthma [32], physical inactivity [33], and type 2 diabetes [34]. However, to our knowledge, studies have yet to examine spatial nonlinear machine learning models to study the prevalence of AD dementia distribution. The National Institute of Aging (NIA) Health Disparities Research Framework advocates for multidisciplinary methodologies to achieve health equity in aging research [35]. This framework organized a spectrum of spatial factors, including environmental, sociocultural, and behavioral elements, to address health disparities, identify intervention targets, and elucidate causal pathways to health outcomes in aging populations [35]. Here, we compile various socioeconomic, environmental, and lifestyle variables together with related health outcomes as predictors. Our main objectives of this study are: 1) to examine the spatial variability of the collected variables on AD dementia prevalence at the county level in the US, 2) to compare the performance of GWRF with widely used linear spatial models (GWR) and non-spatial models (RF and OLS), and 3) to identify county-specific most influential variables and rank their importance. The findings can help public health policymakers to develop more relevant interventions to reduce disparities of AD dementia prevalence in the US rather than a one-size-fits-all policy that might not be efficient on a broader scale.

## METHODS

### Data collection and preparation

The estimated prevalence of AD dementia, serving as the outcome variable, was sourced from Dhana et al. (2020) at the county level across the continental US [36]. This data comprises county-specific estimates for adults aged 65 and older, adjusted on demographic characteristics such as age, sex, race/ethnicity, and education, and was computed from cognitive data from population-based studies and the National Center for Health Statistics.

We collected various predictors (n=24) previously shown to be associated with AD dementia based on the article by Livingston et al. (2020) [37], Alhasan et al. (2024) [23], and domain knowledge. These predictors were categorized into four main risk factor domains: socioeconomic, environmental, lifestyle, and other health outcomes, all at the county level (n=3,032) in the continental US (Table 1).

**Table 1**. Predictors used in this study.

Variables that describe vulnerable communities before, during, and after disasters were developed by the Center for Disease Control and Prevention (CDC) as the social vulnerability index (SVI) [38]. The SVI is constructed using county-level variables obtained from the American Community Survey (ACS) 2016-2020. The estimated rates per county were included for the following SVI variables: below 150% poverty, unemployed, housing cost-burdened, no health insurance, mobile homes, occupied housing with more people than rooms, households with no vehicle, and households without a computer with internet access. More comprehensive descriptions of variables are available from the CDC/ATSDR SVI 2020 Documentation [39]. Additionally, limited access to healthy foods was sourced from County Health Rankings and Roadmaps [40]. To assess the impact of air pollution on AD dementia prevalence, annual average estimates of outdoor concentrations of air pollutants such as O_3_, CO, NO_2_, and PM2.5 were used. These data were estimated using a land use regression model and retrieved from the Center for Air, Climate, and Energy Solutions (CACES) through a partnership with the Environmental Protection Agency [41]. Moreover, remotely sensed normalized difference vegetation index (NDVI) was obtained from Moderate Resolution Imaging Spectroradiometer (MODIS) satellite imagery to represent vegetation density on the ground with a 500 m spatial resolution. NDVI values are calculated from Red (R) and Near Infrared (NIR) spectral bands as (NIR-R)/(NIR+R), ranging from -1 to +1, with higher values indicating denser vegetation cover [42]. Zonal statistics function in ArcGIS Pro 3.3 (ESRI, Redlands, CA) Spatial Analyst Extension was used to compute the average NDVI per county across the continental US.

The age-adjusted prevalence of lifestyle variables per county was extracted from the CDC PLACES dataset, derived from the Behavioral Risk Factor Surveillance System (BRFSS), using a validated method for small-area estimation [43, 44]. BRFSS is the primary system for health-related telephone surveys nationwide, gathering information from US residents about their health behaviors and chronic conditions [45]. The lifestyle variables encompassed the age-adjusted prevalence rates of binge drinking, current smoking, and lack of leisure-time physical activity. Additionally, the age-adjusted prevalence rates for various health outcomes, including depression, obesity, cognitive disability, diagnosed diabetes, hearing disability, and high blood pressure, were sourced from the same repository.

All collected data were compiled at the county level across the continental US and were joined to the administrative boundary shapefile of counties sourced from the US Census Bureau’s TIGER/Line repository [46]. Counties with several missing variables were removed from the geodatabase, resulting in a final set of n=3,032 counties. The linear correlations between the predictors were examined, and the highly correlated variables (|r| > 0.8) and those with high multicollinearity (i.e., variance inflation factor (VIF) > 7.5) were excluded [47]. The final compiled geodatabase was projected to the USA Contiguous Albers Equal Area Conic projection, and the coordinates of the centroid for each county were added to the dataset using the Calculate Geometry function in ArcGIS Pro 3.3 for further geospatial modeling.

### Linear Models

The baseline OLS model was employed to investigate the relationship between AD dementia prevalence and the predictors. The OLS model is specified as follows [48]:

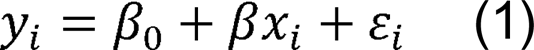

Where *y_i_* represents the prevalence of AD dementia in county *i*; *β*_0_ is the intercept; *β* denotes the estimated coefficients of the predictors; *x_i_* includes the selected predictors; and *ε_i_* is the error term for county *i*. This model has several assumptions, such as spatial stationarity across the study area, linear relationships, independence of observations, normality, random distribution of errors, and is sensitive to outliers. However, spatial dependencies can violate independence and randomness assumptions in practice [49].

To address spatial heterogeneity, geographically weighted regression (GWR), an extension of OLS, was utilized [50]. GWR allows the relationships to vary by location and is expressed as [51]:

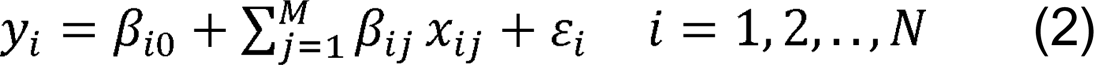

Where *y_i_* represents the *N* × 1 vector of the prevalence of AD dementia in county *i*; *β*_*i*0_ is the intercept for county *i*; *β*_*ij*_ denotes the estimated coefficient for the *j*th predictor *x_ij_* is the *j*th predictor in county *i*; *M* is the number of predictors, *N* is the number of counties, and *ε_i_* is the error term for county *i*. The coefficients for each predictor were estimated as:

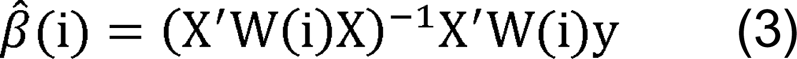

Where *β^^^*(i) represents an *M* × 1 vector of coefficient estimates, *X* is an *N* × *M* matrix of predictors, *W*(i) is an *N* × *N* diagonal weights matrix, y is an *N*× 1 vector of the dependent variable. To compute *W*(i), a kernel function and a bandwidth are specified. The kernel function assigns spatial weights to observations based on their proximity, with higher weights to observations closer to county *i*. The bandwidth controls the level of smoothing; a larger bandwidth incorporates more observations into the estimation. We selected the optimal bandwidth (ranging from 30 to 1000 neighbors) based on minimizing the corrected Akaike Information Criterion [52].

Mapping the locally estimated coefficients can help reveal hidden spatial patterns and the local influence of predictors on AD dementia prevalence [53]. Nevertheless, GWR retains assumptions of local linearity and independence of observations and remains sensitive to outliers [26]. Additionally, local multicollinearity can lead to instability in the parameter estimates [52].

## Nonlinear Models

### Random Forest (RF)

In contrast to linear models, RF and its variants do not depend on specific relationship assumptions for the variables [28], making them well-suited for capturing nonlinear relationships and interactions between variables [54]. RF is a non-parametric machine learning algorithm used for both classification and regression tasks [28, 55, 56]. We divided the entire dataset into two portions: a training set (comprising 2/3 of the data) and an out-of-bag (OOB) set (comprising 1/3 of the data). Using the training set, numerous unpruned regression trees were constructed through bootstrap sampling (i.e., random sampling with replacement) and the random selection of a subset of variables [57]. The prediction accuracy was computed for each tree, and the final output was the average prediction from all regression trees [58].

The OOB data, which was not used during the training process, were used for two primary purposes: 1) evaluating the goodness of fit and overall model accuracy using metrics such as mean square error (MSE), coefficient of determination (R^2^), and mean absolute error (MAE) [59], and 2) determining the relative importance of each variable [27]. To assess variable importance, the values of each variable in the OOB set were randomly permuted, and the OOB error was recalculated. The variable was deemed important if the OOB MSE error increases (%IncMSE) with the permuted values. The greater the increase, the more influential the variable is in predicting AD dementia prevalence [60]. A detailed description of the RF is provided elsewhere [28, 57, 61].

### Geographically Weighted Random Forest (GWRF)

Although RF addresses the limitations of linearity assumptions and susceptibility to outliers in GWR, it remains a global model and fails to capture spatial heterogeneity [62]. The GWRF has recently been developed to overcome the stationary assumption of RF and the limitations of the GWR model [63]. Similar to GWR, GWRF involves locally calibrated models rather than a single global model. GWRF addresses spatial heterogeneity by computing local RF using nearest neighbors for each location based on the spatial weight matrix [64]. An adaptive kernel was used to identify an optimal number of nearest neighbors due to various sizes of counties and uneven distribution of centroid of counties in the US [60]. Detailed discussion on GWRF can be found in [63]. To enhance GWRF interpretability, permutation feature importance was used to rank the local variable importance for each county based on the %IncMSE. Further, we mapped the local variable importance to better understand how, where, and to what extent each variable affects AD dementia prevalence geographically [27]. Moreover, we estimated and mapped the local residuals and local goodness-of-fit statistics for training and out-of-bag (OOB) samples to assess the model performance in different regions.

### Hyperparameter Tuning

RF and GWRF are less susceptible to overfitting due to the creation of many slightly different regression trees [28, 65]. They are robust prediction models when the hyperparameters are appropriately tuned, especially with large datasets [28]. To minimize overfitting, the hyperparameters of the RF were fine-tuned using the random grid search, optimizing the number of variables randomly sampled (mtry) and the number of trees (ntree) on a large dataset (n=3,032) [27, 34]. A 10-fold cross-validation method was used to determine the optimal hyper-parameters from all possible combinations. We also used these hyperparameters for GWRF. Moreover, the optimal bandwidth (maximum number of neighbors to calibrate local models) in GWRF was selected based on minimized OOB error [63].

We applied the Getis-Ord Gi* statistics to assess the clustering of high (hotspots) or low (cold spots) residual values within the entire dataset [66]:

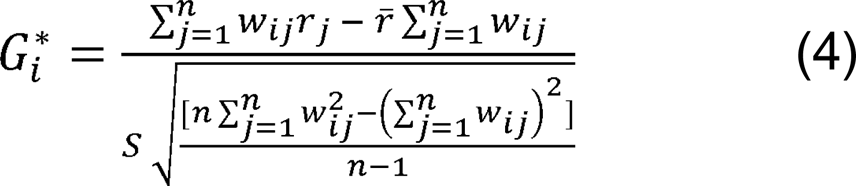

Where *r*_*j*_ is the residual at county *j*; *W_ij_* is the spatial weight between county *i* and county *j*, which is non-zero for counties that share borders; *r̄* is the mean of residuals; *S* is the standard deviation of residuals. A positive *G_i_^*^* value above the expected indicates a hotspot, while a negative value below the expected indicates a cold spot [67]. All statistical analyses were conducted using randomForest, SpatialML, and CARET packages in R, with mapping performed in ArcGIS Pro 3.3.

## RESULTS

Preliminary descriptive statistics showed that the AD dementia prevalence (%) in the continental US ranged from 5.6% in Lovington County, Texas, to 18.4% in Presidio County, Texas. The mean prevalence rate (adjusted for age, sex, race/ethnicity, and education) was 11.17%, with a standard deviation of 1.43%. The highest rates were predominantly found in southern states, including North Carolina, South Carolina, Georgia, (southern) Florida, Tennessee, Alabama, New Mexico, and (southern) Texas (Fig 1). The states with the lowest (mean) prevalence rates, respectively, were Utah (9.70%), Vermont (9.76%), and Idaho (9.87%), while those with the highest (mean) rates were Mississippi (13.10%), South Carolina (12.46%), and Louisiana (12.42%). Among the 24 initial predictors, four variables (i.e., below 150% poverty, the age-adjusted prevalence of cognitive disability, diagnosed diabetes, and high blood pressure) were removed due to a high correlation with other variables or high VIF. The correlation coefficients matrix for all predictors is available in the Supplementary File. This resulted in 20 final predictors, with all correlation coefficients below the threshold value of |0.8| and VIFs less than 7.5, suggesting low multicollinearity. Overall, nonlinear models (i.e., RF and GWRF) demonstrated better fit and accuracy than linear models (i.e., OLS and GWR). In terms of R² for the training dataset, the OLS model had the worst performance (R² = 60.6%, MAE = 0.68, RMSE = 0.89), while the GWRF model had the best performance, with an R² close to 1 and the slightest error (MAE = 0.04, RMSE = 0.05). However, overfitting using the training dataset for nonlinear models is likely. Table 2 presents the evaluation metrics for both the training and OOB datasets. To avoid overfitting before fitting the RF and GWRF models, we tested various combinations of hyper-parameter values through 10-fold cross-validation. The optimal hyperparameters were based on the adaptive kernel, bandwidth = 220 nearest neighbors, ntree = 5,000, and mtry = 10. For comparison, these parameters were fixed for both the training and OOB datasets of RF and GWRF models. Based on the OOB accuracy assessment, both models performed well, with GWRF demonstrating slightly better performance, showing a 4.0% increase in R^2^ (Table 2). Additionally, due to the local implications and interpretations of GWRF, such as spatially varying relationships, local feature importance, flexible bandwidth selection, and integration with GIS for visualization and targeted decision-making, GWRF was used for further inferences.

**Fig 1.**
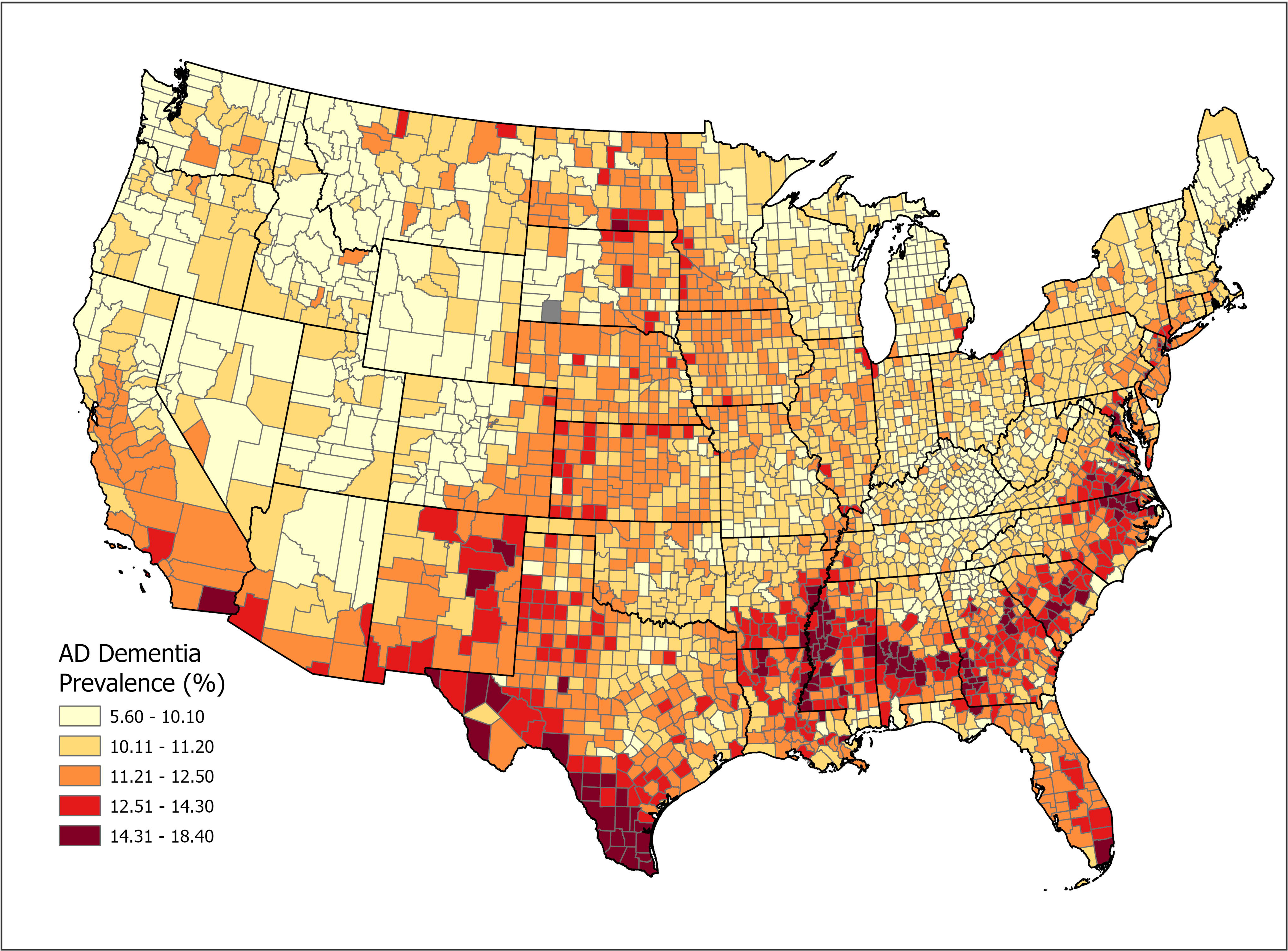
Spatial distribution of AD dementia prevalence (%) in the continental US.

**Table 2**. Comparative performance of the models for AD dementia prevalence rates in the US.

According to RF permutation-based feature importance, the variable “Depression” was the most significant contributor (highest %IncMSE), followed by “Physical Inactivity”, “Obesity”, “PM2.5”, “Households No Vehicle”, “Binge Drinking”, and “Smoking”. However, “Household Crowded” was the least contributing variable to AD dementia prevalence. Figure 2 depicts the relative importance of variables to AD dementia prevalence based on the RF model.

**Fig 2.**
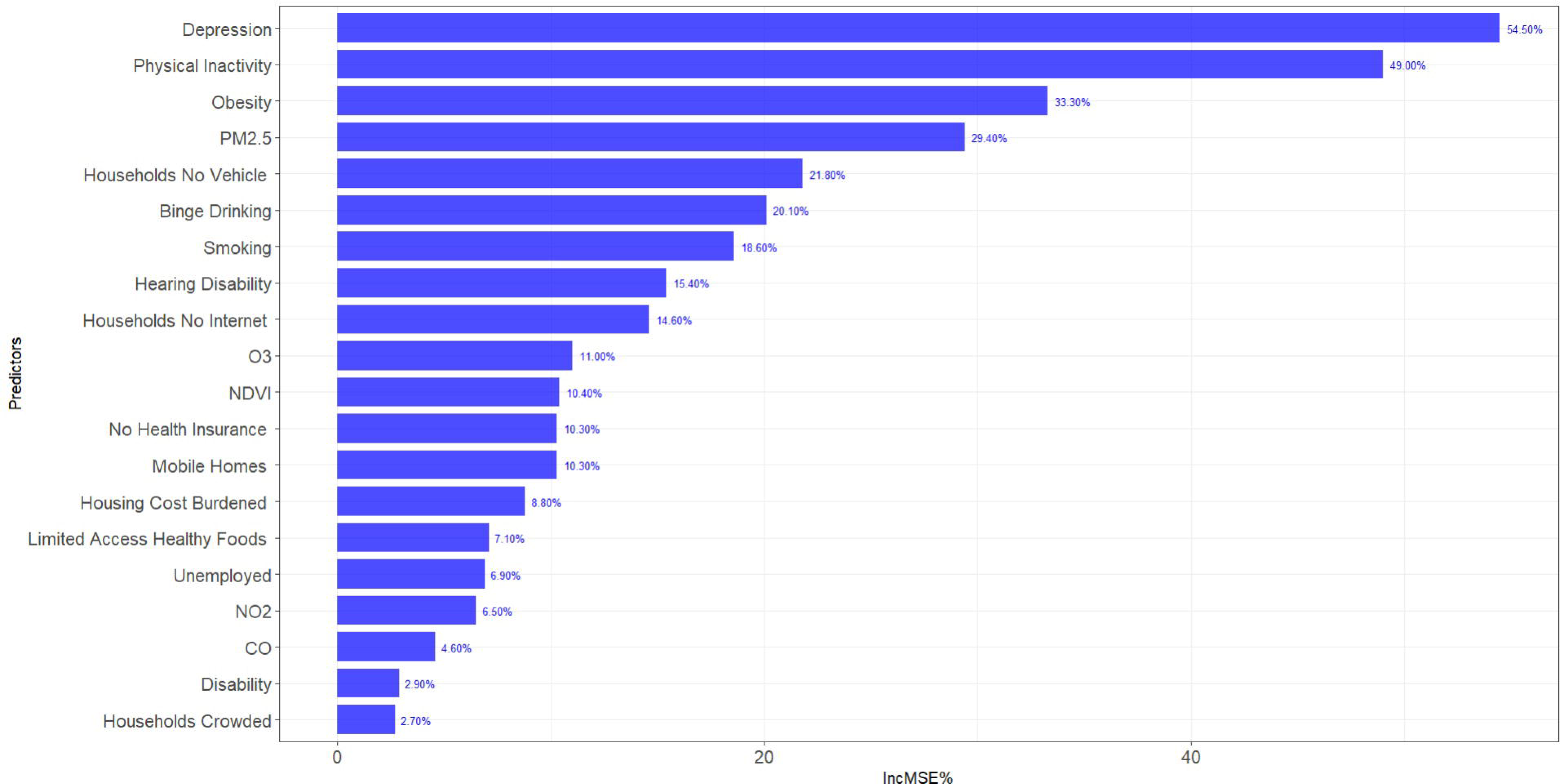
The relative importance of variables to AD dementia prevalence using permutation-based feature importance. A higher increase in MSE (%IncMSE) corresponds to higher importance.

The percentage of “NDVI” was the leading primary factor for the prevalence rate of AD dementia in most US counties (n= 458, 15.1%), especially in the northern and central states such as Minnesota, Wisconsin, and Missouri. “Mobile Homes” was the most influential factor in (n=404, 13.3%) of US counties, notably in Ohio, Kentucky, North Dakota, and South Dakota. “Physical inactivity” was the primary factor in (n=390, 12.7%) of south and southwestern counties, predominantly in Arizona, New Mexico, Colorado, Utah, and southern Texas. “Depression” was the main influential factor in (n=348, 11.5%) of US counties, particularly in most counties of Virginia, West Virginia, Maryland, and North Carolina. Figure 3 illustrates the geospatial distribution of local primary factors of AD dementia prevalence across the US and the corresponding proportion of counties.

**Fig 3.**
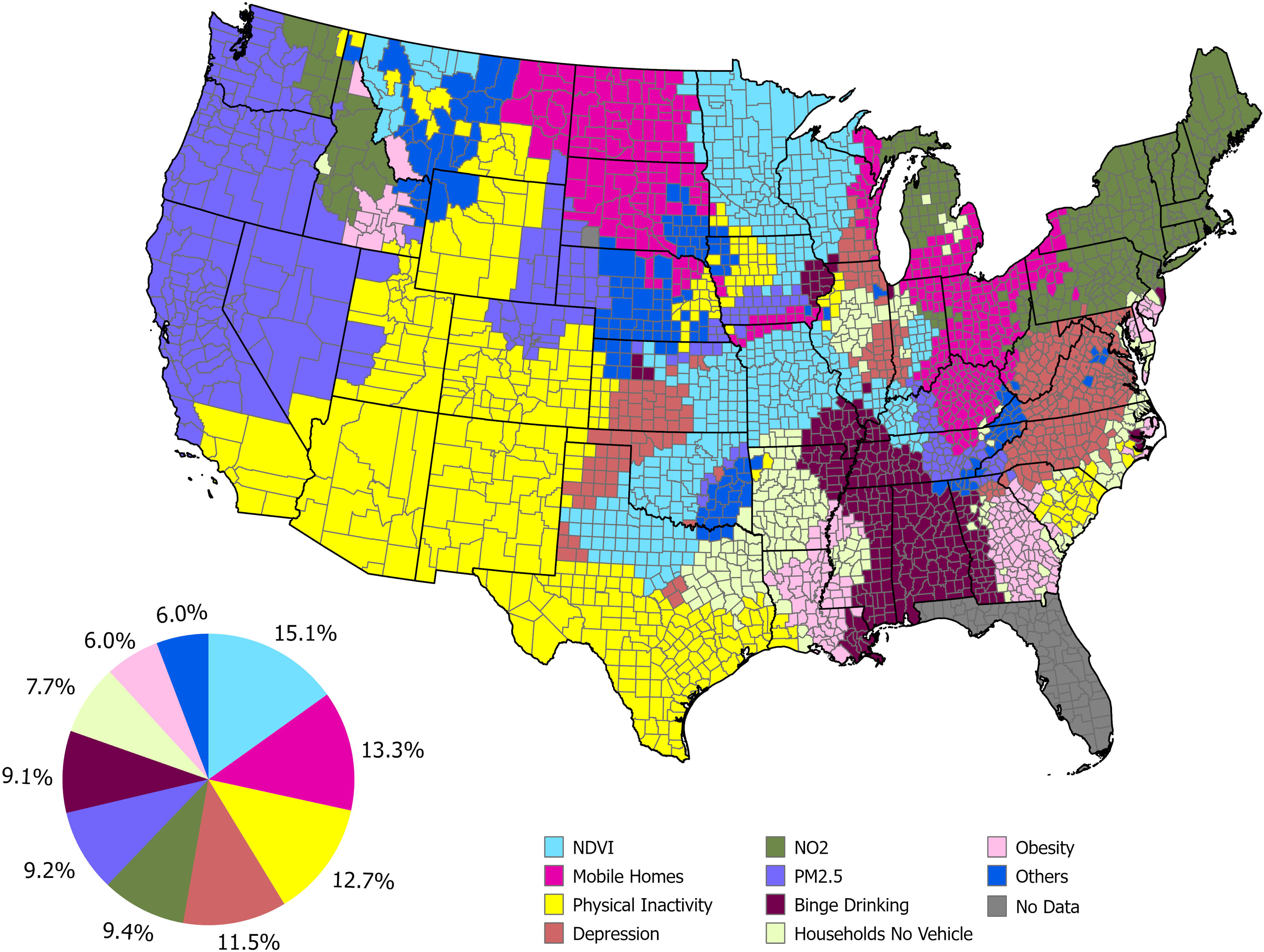
Spatial distribution of primary local factors influencing AD dementia prevalence in the US.

The local effects of the top important variables were mapped, regardless of their primary status. ‘NDVI’ had the largest and most widespread impact in northern and central states, with a small cluster of counties in Texas, while having the lowest impact in the eastern and western regions. ‘Mobile Homes’ significantly influenced AD dementia prevalence in the northern US, particularly in North Dakota, South Dakota, Pennsylvania, and New York, with the least impact in the south, northwest, and central regions. ‘Physical inactivity’ had the highest impact in the southern half of the US, notably in New Mexico, Arizona, Colorado, Louisiana, Mississippi, and South Carolina. ‘Depression’ most affected the southern states and had the least impact on the northern states. ‘NO_2_’ was most significant in the northeastern states, including Maine, Vermont, New Hampshire, Massachusetts, New York, Connecticut, and Rhode Island, while it had a low impact almost everywhere else. ‘PM 2.5’ had the most significant effects in the western half of the US, especially in northern Colorado, Wyoming, Arizona, and Nevada. ‘Binge drinking’ and ‘No vehicle’ exhibited nearly identical geographic patterns, with the highest impacts in southern states, particularly Alabama and Missouri, and the lowest impacts in the western and northern US. Figure 4 illustrates the local impact of the top variables on AD dementia prevalence in the US.

**Fig 4.**
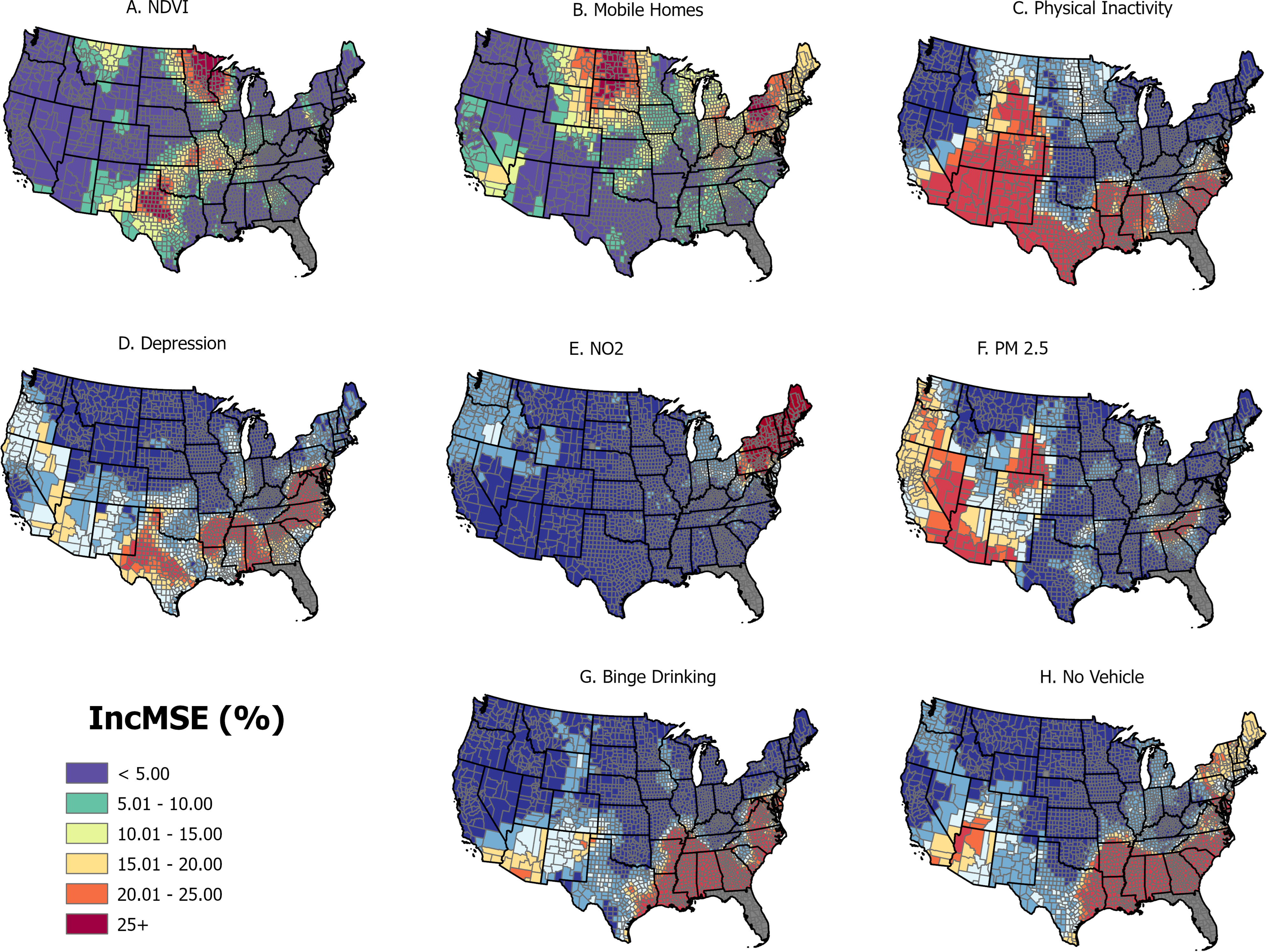
The local impact of the top variables on AD dementia prevalence in the US.

The local R² of the GWRF model in explaining the variance of AD dementia prevalence ranged from 0.04 to 0.84 (median = 0.51, standard deviation = 0.16). The model tended to perform better in most counties in the southern, eastern, and northeastern regions (local R² > 0.7), while it underperformed in the central and northwestern US (local R² < 0.2). This indicates that some key variables were likely missing in these areas. Figure 5A illustrates the local R² of the GWRF model. The Getis-Ord Gi* analysis showed that the residuals of the GWRF model were mainly randomly distributed, with a few hotspots or cold spots indicating areas of overprediction or underprediction (Figure 5B).

**Fig 5.**
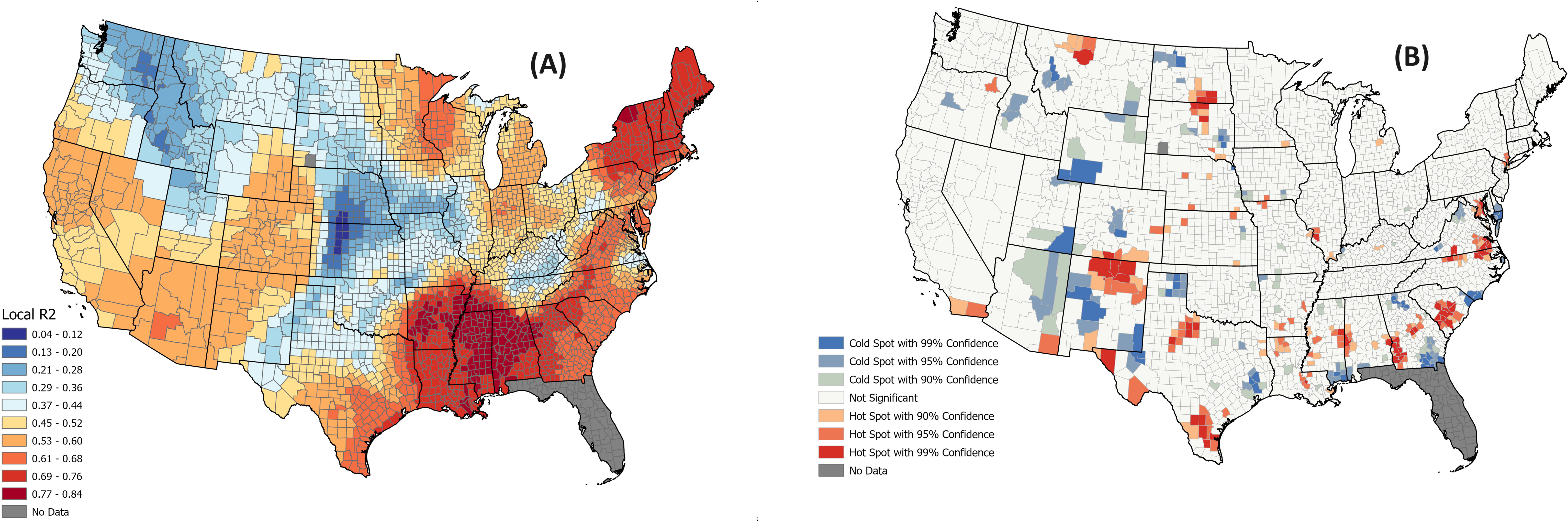
The geospatial distribution of A) Local R^2^ of the GWRF model and B) Getis-Ord Gi* hotspots/cold spots of residuals.

## DISCUSSION

Inspired by the NIA’s Health Disparities framework [35] and leveraging the foundational work by Livingston et al. (2020) [37], we compiled a geodatabase of socioeconomic, environmental, and lifestyle variables along with other pertinent health outcomes at the county level in the continental US. Utilizing advanced spatial technologies such as GIS, remote sensing, spatial statistics, and machine learning model, we conducted a geospatial machine learning modeling of the distribution of AD dementia prevalence to better understand the local impacts of these variables in the continental US. The results of our study can guide interventions such as enhancing green spaces to improve cognitive health, reducing air pollution, and promoting physical activity through initiatives, for instance, for mobile home communities. The findings of this study can serve as the foundational baseline to support the development of cost-effective, place-based interventions and more refined policy development across the nation. We sourced the AD dementia prevalence data from the study by Dhana et al. (2023) [36]. The advantage of using this data, as compared to conventional estimates relying on medical reports, claims, and national surveys, is its mitigation of significant underestimation, particularly among racial and ethnic minority groups. Conventional methods often miss cases due to discrepancies between data sources and systemic diagnostic biases. In contrast, the data used in this study incorporated findings from a diverse population-based study and adjusted for demographic variations (i.e., age, sex, race/ethnicity, and education) at the county level, providing a more accurate assessment of the factors impacting AD dementia prevalence in the US [36]. African American and Hispanic older adults face a potentially 2-fold higher risk of AD dementia [68]. Therefore, the involvement of racial and ethnic minority groups in AD clinical research is emphasized [69, 70].

Geospatial modeling of AD dementia remains relatively sparse, with conventional geospatial linear models like GWR constrained by local linearity, sensitivity to outliers, and multicollinearity [26, 27]. Thus, our study adopted a nonlinear spatial approach to circumvent these limitations while incorporating variable interactions, resulting in superior performance to conventional linear models. Our findings confirmed and extended previous research on other health outcomes using GWRF [32, 71]. Although the model exhibited proper performance in most counties, it showed low goodness-of-fit in central and northwest US counties, indicating the necessity for integrating additional critical variables and enhancing data quality. Notably, while OOB error assessment for the GWRF was marginally better than RF, it provided insights into local feature importance, local goodness of fit, and local effects of the predictors.

The permutation-based feature importance analysis of GWRF suggested that among environmental factors, NDVI was found to be the most important factor in 15.1% of US counties. Counties most influenced by this factor lie in the northern and central regions. Numerous studies have linked NDVI, or neighborhood greenness, to improved cognitive function and MRI results [12]. A 13-year cohort study involving over 1.1 million older adults from 2001 to 2014 indicated that an increase of one interquartile range in surrounding greenness was associated with a 4–5% decrease in premature mortality from all neurodegenerative diseases, including AD [72]. However, one study found that this association is only significant for those with low genetic risk [73]. NO2 and PM 2.5 emerged as the other important environmental variables and the primary factors in 9.4% and 9.2% of US counties, notably in northeastern and western states, respectively. This concurs with previous studies associating air pollution with cognitive decline [74, 75]. PM 2.5, typically composed of heavy metals such as nickel, lead, cadmium, arsenic as well as organic carbon, can easily cross the blood-brain barrier and reach the central nervous system [76, 77]. They can induce systemic inflammation, cell death, and DNA damage in the human brain by producing reactive oxygen species, which are critical for developing AD dementia [78, 79].

Lack of leisure-time physical activity was identified as the most important modifiable lifestyle variable in explaining AD dementia prevalence. It was found as the primary variable in 12.7% of US counties, particularly affecting New Mexico, Arizona, Colorado, Louisiana, Mississippi, and South Carolina. A review article of various study designs indicated that higher levels of physical activity are linked to a lower risk of late-life dementia. Physical activity can enlarge prefrontal and hippocampal brain areas, potentially mitigating memory impairments [80]. Possible reasons for the regional impact might include access to recreational facilities, cultural attitudes toward physical activity, and environmental factors that either promote or hinder an active lifestyle. For instance, a cross-sectional analysis of data from 50,884 women revealed that those residing in areas with the highest exposure to greenness were more inclined to participate in higher levels of physical activity (more than 67.1 MET hours per week) compared to those in areas with the lowest level of greenness [81]. Binge drinking was the second most important lifestyle variable that impacted AD dementia prevalence and the primary important variable in 9.1% of US counties. A 25-year follow-up analysis among a cohort of 554 Finnish twins aged 65 years and older at the time of assessment revealed that midlife binge drinking was associated with a relative risk of 3.2 for dementia [82]. Binge drinking can markedly worsen cognitive deficits [83]. A review study investigated how binge drinking is associated with the development of cognitive impairment among young adults in the UK. They found deficits in brain regions such as the frontal lobe, temporal lobe, and hippocampus. However, the studies lacked sufficient evidence to generalize the findings [84].

Among the socioeconomic factors, “living in mobile homes” and “households with no vehicle” were identified as the most significant variables impacting the prevalence of Alzheimer’s disease (AD) dementia. These factors were primary variables in 13.3% and 7.7% of US counties, respectively. While the exact mechanisms linking mobile homes to AD dementia prevalence are unclear, possible explanations include the increased socioeconomic stress faced by residents due to financial instability, limited healthcare access, and increased exposure to air pollution [85], all of which can contribute to a higher risk of cognitive decline. Additionally, no vehicle can exacerbate isolation and restrict access to healthcare services and social activities, all of which are crucial for maintaining cognitive health, thereby increasing the prevalence of AD dementia. While this study offers valuable insights, it does have its limitations. The data utilized in this study were primarily based on model-derived estimations of AD dementia prevalence and telephone survey responses from BRFSS, which are susceptible to recall and social desirability biases. The AD prevalence data obtained from [36] focused on three major racial/ethnic groups (Black/African American, Hispanic, and White) and simply assumed that other races (i.e., Asian American and American Indian/Alaska Native individuals) have a similar risk to White individuals. Additionally, due to the ecological fallacy, the conclusions cannot be extended to the sub-county or individual levels and should only be interpreted at the county level. Another limitation that might reduce the statistical power of the analysis is the absence of various key variables in certain states, notably Florida, and particularly Alaska and Hawaii, where each has a racial and ethnic minority group of approximately 40%. This led to the exclusion of these states from the analysis. Moreover, it should be acknowledged that the feature importance technique used in this study can be impacted by inter-feature correlation and multicollinearity, which can lead to misleading importance scores as permuting one feature may not accurately reflect its individual impact due to the correlated variables.

Although we included a selection of well-recognized variables, future research should incorporate distinctions between rural and urban settings and consider the built environment and social isolation variables. Additionally, conducting research at finer spatial scales while maintaining a national scope is crucial for more targeted interventions. Subsequent research should involve spatial causal inference and mediation analyses to identify mediators and mechanisms.

In conclusion, our findings underscore the considerable spatial variability in the factors associated with AD dementia prevalence across US counties. This suggests that local and regional governments should implement targeted interventions based on specific regional variables. For example, some areas may benefit from prioritizing improving green space and reducing air pollution, while others might focus on modifying lifestyle variables such as increasing physical activity and reducing binge drinking. Moreover, the fit of the local model highlights the urgent need for more comprehensive data collection and analysis to better understand how disparities impact AD dementia prevalence. Together, these insights can pave the way for more nuanced research and targeted interventions to mitigate the prevalence of AD dementia in the US.

## Data Availability

All data produced in the present work are contained in the manuscript

## ACKNOWLEDGMENTS

None

## CONSENT STATEMENT

No human subjects participated in this study, consent was not necessary.

---

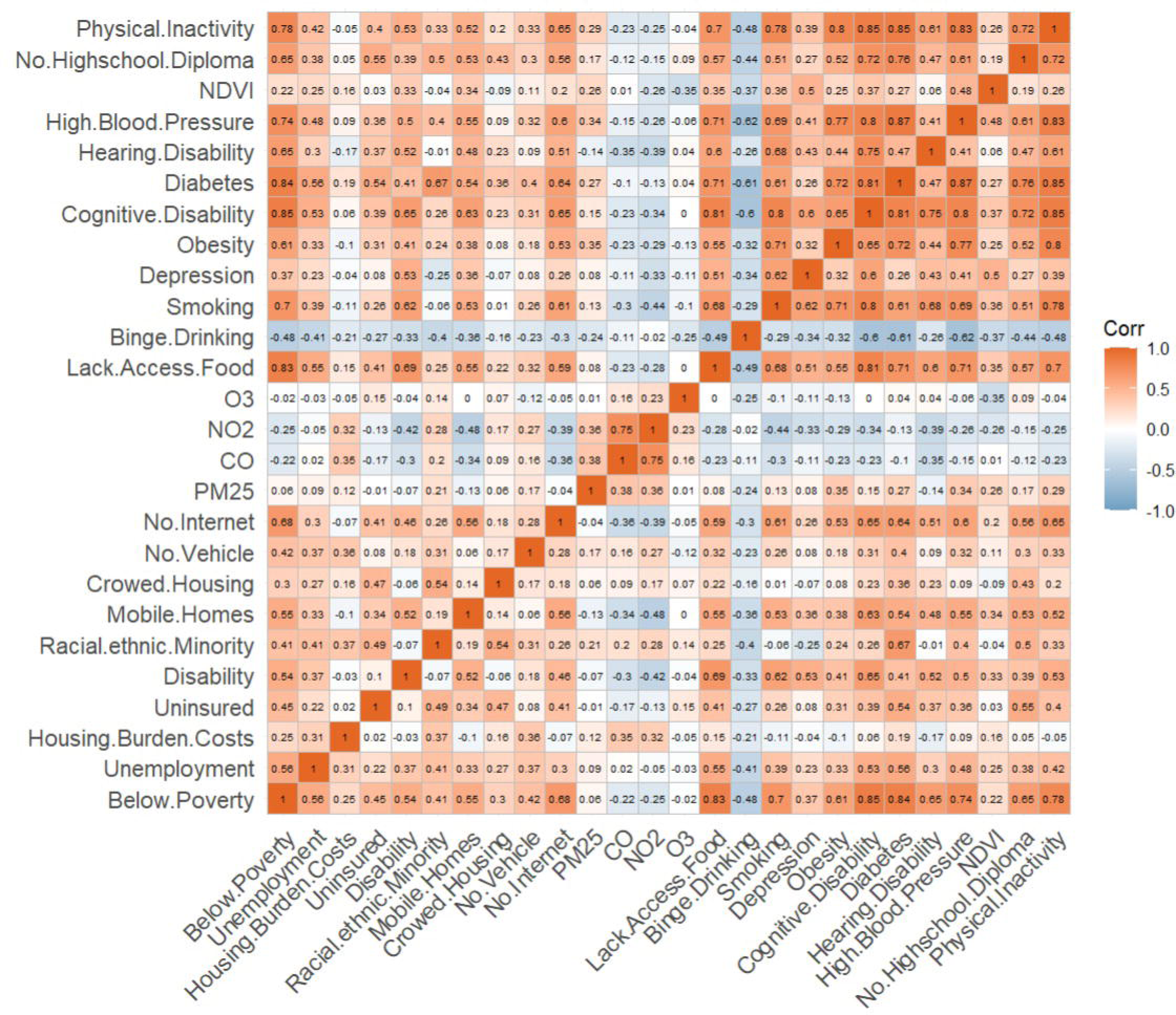

